# PANCDetect: Early Detection of Pancreatic Cancer from Multimodal EHR data with LLM Embeddings

**DOI:** 10.1101/2025.10.03.25337210

**Authors:** Zicheng Jin, Xuhui Guo, Zehua Wang, Qiang Yang, Xiaotong Yang, Xinyu Zhang, Rui Yin, Lana X. Garmire

## Abstract

**Background:** Pancreatic cancer (PANC) is often diagnosed at late stages due to the absence of specific early symptoms, resulting in one of the highest cancer mortality rates. While imaging modalities such as MRI and CT offer high diagnostic accuracy, their population-wide application is however impractical due to the cost. Electronic health records (EHRs) provide a routine, easily accessible, longitudinal and scalable data source for risk prediction, particularly for diseases with no specific symptom such as PANC.

**Method:** We introduce PANCDetect, a multimodal framework that leverages large language model (LLM)-derived embeddings of diagnoses, procedures, medications, and laboratory tests, and integrates these data modalities through a Transformer-based architecture. We train the model on MarketScan (≈250M patients), and validate it externally on additional large real-world EHR datasets of University of Michigan Precision Health, or UMPH data (n≈6M patients) and OneFlorida+ data (n≈26M patients). We then fine-tuned the general model on UMPH EHR data. We evaluated the performance of both models using metrics including area under the receiver operating characteristic curve (AUROC) and area under the precision-recall-gain curve (AUPRG). We assessed the top predictive features with integrated gradients (IG).

**Result:** In the MarketScan cohort, PANCDetect achieved an AUROC of 0.812 and AUPRG of 0.851 at the 6-month prediction window, and an AUROC of 0.735 and AUPRG of 0.629 for 60-month prediction, significantly outperforming CancerRiskNet. External validation on UMPH and OneFlorida+ demonstrated good generalizability, with 6-months AUROC scores of 0.711 and 0.793, respectively. Fine-tuning on UMPH with laboratory data further improved performance, reaching an AUROC of 0.927 and an AUPRG of 0.979 at 6 months. Even at the 60-month horizon, the refined PANCDetect model maintained strong performance, with an AUROC of 0.835 and AUPRG of 0.911. Attribution analysis highlighted type 2 diabetes, pancreatic diseases, personal and family cancer history as the most important risk factors.

**Conclusion:** PANCDetect is the state-of-the-art method integrating multimodal EHR data with LLM embeddings for accurate, interpretable, and generalizable early prediction of pancreatic cancer. This framework holds promise for precision screening of high-risk patients, with the potential to improve survival outcomes without increasing healthcare costs.

## Introduction

Pancreatic cancer (PANC), is characterized by the lack of specific early clinical manifestations.^1^ In many cases, individuals may experience vague or nonspecific symptoms that precede diagnosis by years.^2^ Based on a global analysis of data from 48 countries, incidence and mortality of PANC are rising globally, particularly among women and individuals aged 50 and older, with concerning increases also seen in younger populations.^3^ The absence of validated screening protocols, lack of reliable early biomarkers, combined with the retroperitoneal and anatomically inaccessible location of the pancreas, hinders timely detection. As a result, less than 20% patients are diagnosed at an early, resectable stage, resulting in a persistently high mortality rate of this malignancy.^4^ Additionally, delayed diagnosis of PANC imposes substantial financial burdens on patients, families, and healthcare systems, as late-stage treatments are far more costly than early surgery with adjuvant therapy.^5^

Routine MRI and CT can be an effective screening method for detecting PANC with high sensitivity and specificity,^6^ but applying cross-sectional imaging at a population scale is impractical due to prohibitive costs and resource burden.^7^ Electronic health record (EHR) is a much more economical, accessible and scalable data resource. EHR data include valuable baseline information such as demographics, as well as the longitudinal information including disease diagnoses, laboratory results, medication, and procedures, making them well-suited for early detection and prognosis modeling.^8^ Moreover, the standardized EHR frameworks allow them to be more AI-ready, and comparable across different healthcare systems, which enhances the generalizability of predictive models across health centers.^9–11^

Previous transformer-based pretrained models demonstrated the feasibility of applying self-attention architectures to structured EHR data for the general prediction of future diagnoses of patients. Models such as BEHRT^12^, Hi-BEHRT^13^, and Med-BERT^14^ adapted BERT-style encoders to longitudinal claims and clinical coding sequences, effectively capturing temporal dynamics, disease progression, and patient heterogeneity. Building on this trend, researchers have applied Transformers to pancreatic cancer risk prediction, leveraging self-attention mechanisms to better model temporal dependencies and disease evolution. Moreover, recent advances in large language models (LLMs) offer new opportunities to extract clinically meaningful patterns hidden among the millions of patients’ EHR data. By treating EHRs as sequences of “clinical tokens”, such as diagnoses (ICD/SNOMED), procedures (CPT), medications (RxNorm/NDC), and discretized lab values, pretrained LLMs can learn rich contextualized embeddings that capture clinical semantics and longitudinal continuity across care settings.^15,16^ General-purpose LLMs such as GPT have been adapted to biomedical corpora (e.g., BioGPT^17^) and even to institution-scale EHR data (e.g., NYUTron^18^), enabling predictive modeling, free-text understanding, and zero or few-shot adaptation to diverse downstream tasks.

A few recent studies have leveraged EHR data to build models tailored for predicting PANC risk, enabling identification of high-risk patient cohorts^19–22^. Among these efforts, CancerRiskNet^21^ stands out as a representative study that demonstrated the feasibility of using Transformers in this domain. However, its multi-window prediction design, which does not exclude disease events occurring shortly before cancer diagnosis, carries the risk of data leakage, potentially inflating accuracy estimates. Moreover, CancerRiskNet relies solely on diagnosis codes, and it is acknowledged that incorporating other clinically informative modalities could further improve prediction accuracy with the real-world availability of data. Many existing models are trained on relatively small datasets, restricting generalizability across diverse populations and health systems^22^. To address these limitations and take advantage of LLMs, we propose PANCDetect, a Transformer-based multimodal prediction framework trained on the MarketScan dataset, which contains over 110,000 pancreatic cancer patients, making it the largest resource of its kind to date (Fig. 1). PANCDetect not only incorporates diagnostic information but also procedures, medications, and laboratory tests into LLM-based embeddings. We also employed a more rigorous exclusion strategy to minimize data leakage in modeling and improve reliability. To validate the generalizability of our approach, we tested and fine-tuned the model across two independent healthcare systems. Consequently, PANCDetect substantially enhances prediction accuracy and clinical applicability for early pancreatic cancer detection as compared to other earlier efforts.

**Fig 1.**
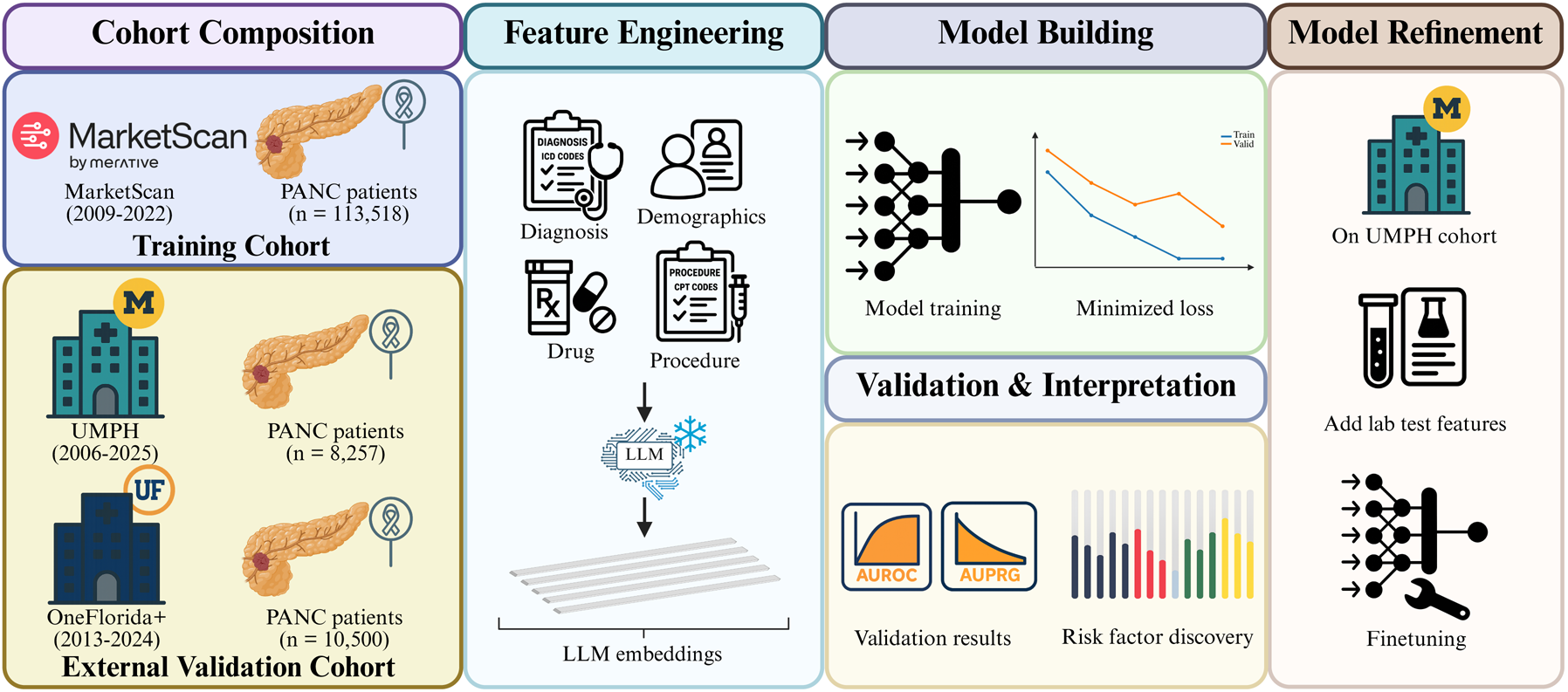
Overall workflow of this study. (1) Cohort selection: The MarketScan cohort was used for model training, while two independent cohorts (UMPH and OneFlorida+) served as external validation datasets. (2) Feature Engineering: During model training, diagnoses, procedures, and medications were extracted as inputs. Standardized textual descriptions of these medical codes were encoded into embeddings using a pretrained LLM. (3) Model Building and Validation: The generated embeddings were passed into a Transformer-based model, with the best model selected based on minimized validation loss. Evaluation was conducted on the held-out test set and external validation cohorts. Attribution analysis using Integrated Gradients (IG) was applied to identify key risk factors. (4) Model Refinement: Fine-tuning was further performed on the UMPH cohort to include additional laboratory tests into the pretrained model using MarketScan data. UMPH: University of Michigan Precision Health; PANC: pancreatic cancer.

## Methods

### Study Cohorts

This study is conducted on three different retrospective datasets: Merative MarketScan Databases,^23^ University of Michigan Precision Health (UMPH),^24^ and OneFlorida+ Clinical Research Network.^25^ MarketScan is a commercial database, which we have access to through the University of Michigan. UMPH and OneFlorida+ data usage was approved by the Institutional Review Board (IRB) of the University of Michigan Medical School (HUM#00259964) and the IRB of the University of Florida (#IRB202400752), with a waiver of informed consent for secondary use of de-identified EHR data. No data from UMPH or OneFlorida+ was shared outside the respective organizations, ensuring compliance with data security and confidentiality.

MarketScan Database contains records from approximately 250 million patients from 2009– 2022. We identified 113,518 individuals diagnosed with PANC based on the diagnosis codes listed in Supplementary Table 1. UMPH includes approximately six million patients from 2006 to 2025, among whom 8,257 individuals are recorded as PANC.. OneFlorida+ Clinical Research Network encompasses around 26 million patients from 72 hospitals across Florida, Alabama, Georgia, and Arkansas. We identified 10,500 individuals diagnosed with PANC during 2013–2024. For each cohort, we identified corresponding matched control patients, by applying a 1:5 matching strategy based on sex, age, and geographic location (Supplementary Fig. 1) to mitigate the substantial class imbalance.

We then used the MarketScan cohort as the primary training set given its largest PANC patient size. We performed external validation on UMPH and OneFlorida+, and fine-tuned the model on the UMPH cohort for better adaptation to the local population characteristics.

### Feature Embedding

As shown in Fig. 2A, the input features comprise time-varying information and time-invariant features. Time-varying information includes EHR clinical events such as diagnoses, procedures, and medications, each annotated with temporal markers such as visit date and patient age at that visit. Time-invariant attributes include sex and geographic location. For the time-variant features, standardized textual descriptions of medical codes are encoded using the OpenAI text-embedding-3-small model to generate dense semantic representations (without patient private information). These embeddings are combined with positional embeddings to capture temporal order and irregular intervals after linear projection, thereby enhancing the representation of longitudinal patient trajectories. During fine-tuning on the UMPH cohort, we integrated laboratory test results as a new input modality, using the same embedding generation method to maintain consistency while enabling exploration of richer multi-modal inputs in a real-world clinical setting. Other time-invariant features (e.g., sex and geographic location) are mapped into dense representations using a feed-forward layer before being concatenated with the longitudinal sequence representations.

**Fig 2.**
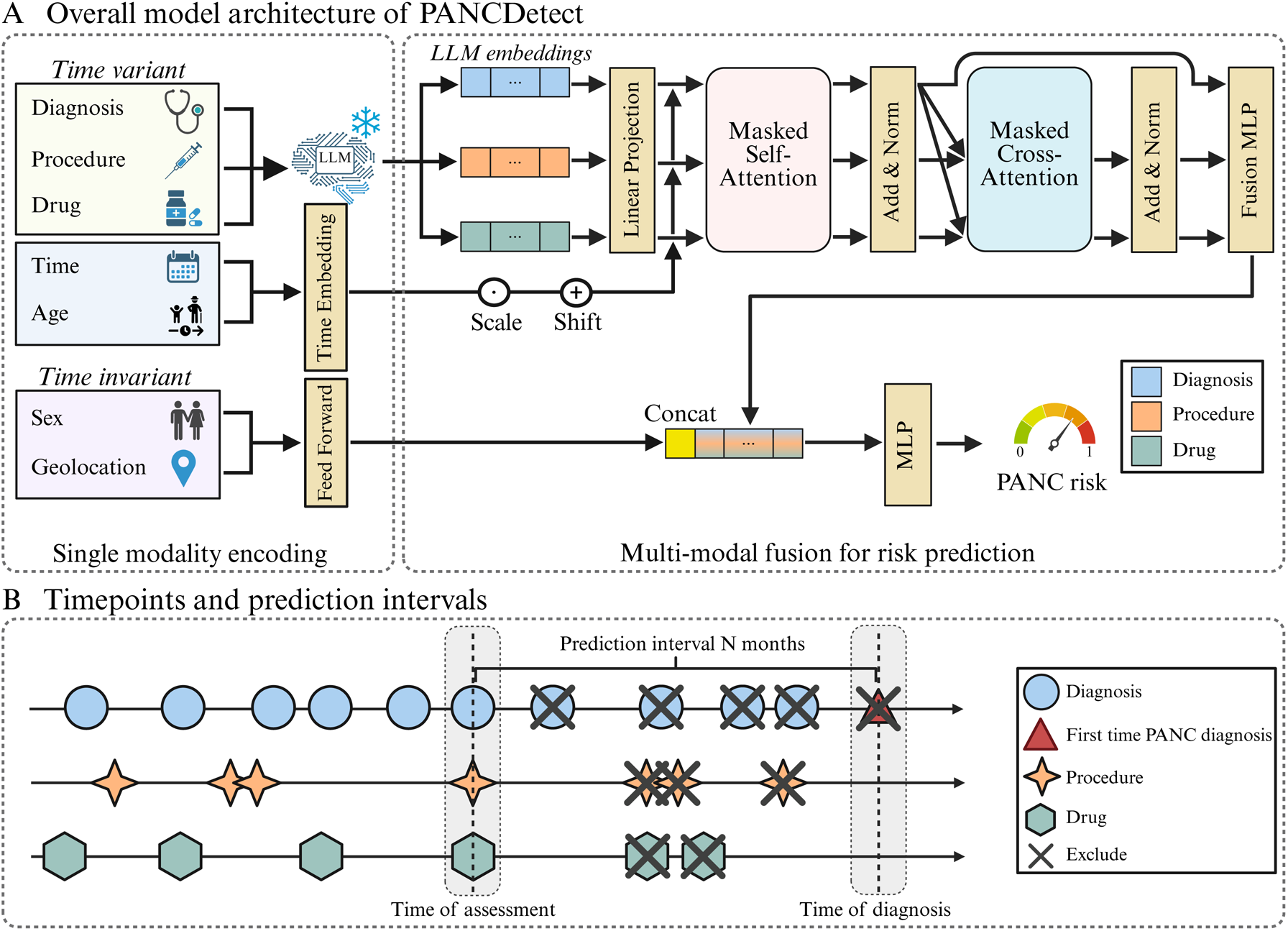
Model structure of PANCDetect and single-interval prediction design. Panel A shows the overall model structure of PANCDetect. Firstly, time-varying EHR events (diagnoses, procedures, medications) are encoded with LLM-based embeddings and temporal markers to represent longitudinal patient trajectories and then processed by a Transformer with masked self- and cross-attention. The resulting sequence representation is pooled into a patient vector and late-fused by concatenation with time-invariant attributes (e.g., sex, geographic location); a final MLP maps the fused vector to a continuous pancreatic cancer risk score. Panel B illustrates the single-interval prediction design, where EHR events within *N* months before cancer diagnosis are masked to reduce data leakage, and trajectories are truncated according to predefined prediction windows (6, 12, 24, 36, and 60 months). LLM: large language model; MLP: multi-layer perceptron; PANC: pancreatic cancer.

### Multi-modal Fusion Strategy

The proposed framework integrates multi-modal information within a Transformer-based architecture (Fig. 2A). Here, three modalities, namely diagnoses, procedures, and medications, are modeled through self-attention blocks to capture intra-modality dependencies. Cross-attention mechanisms are then applied, using diagnosis events as queries and procedure and medication events as key–value inputs, enabling the model to learn complementary information across modalities. During fine-tuning on the UMPH cohort, additional modalities, such as laboratory tests, can be incorporated in the same manner. The resulting multimodal representations are concatenated and passed through a fusion multilayer perceptron (MLP) to construct longitudinal patient representations, which are subsequently combined with time-invariant features to predict pancreatic cancer risk (Fig. 2A). This framework is modular and can be readily extended to accommodate further modalities as they become available, such as in the refined model using UMPH data.

### Diagnosis Prediction Intervals

Compared with prior studies^21,22^, we adopted a single-interval prediction design. When predicting “PANC diagnosis within N months”, all EHR information on visits occurring within the N-month interval before the PANC diagnosis was masked and only earlier EHR information was retained to train the model (Fig. 2B). Specifically, for each PANC patient, we extracted all diagnoses, procedures, and medication records occurring prior to the PANC diagnosis date. We truncated the clinical trajectories according to different predefined prediction intervals N (N=6-, 12-, 24-, 36-, 60-month). For non-PANC controls, the reference date was defined as the end of their observation period, and their records were truncated backward from this time point using the same prediction intervals. This strategy is essential to reduce the risk of data leakage from prodromal symptoms very close to the diagnosis date and provides a more realistic assessment for early detection potential.

### Implementation Details & Evaluation Metrics

Patients with valid longitudinal trajectories after preprocessing are randomly split into training, validation, and test sets in an 8:1:1 ratio. The model was trained using the binary cross-entropy loss with logits, optimized with AdamW (initial learning rate of 10^-4^, weight decay applied). All training experiments were performed on 4 NVIDIA A40 GPUs for 10 epochs with a batch size of 128. Model performance was primarily assessed using several metrics: (1) the area under the receiver operating characteristic curve (AUROC), (2) the area under the precision-recall-gain curve (AUPRG),^26^ normalized adjusted area under the precision–recall curve (AUPRC) with precision-gain and recall-gain scores, which handles the imbalances between case vs control samples. For each metric, we used the 95% confidence interval (CI) estimated via 200 bootstrap resamples. To provide a more comprehensive assessment, we also reported precision, recall, F1 score, and specificity (Supplementary Table 2-5).

### Feature Interpretation

To enhance model interpretability, we applied integrated gradients (IG)^27^ to perform attribution analysis on the input embeddings. IG is a path-integrated, gradient-based attribution method that quantifies feature contributions to the prediction outcome (see Supplementary Appendix for formulation). Specifically, it computes feature attributions by accumulating gradients of the prediction score with respect to the input along a straight-line path in the embedding space, interpolating from a baseline input to the actual input. We defined the baseline as a zero embedding vector, representing the absence of any clinical information. Based on the attribution results, we aggregated importance scores for all inputs and identified the clinical features most relevant to PANC risk prediction.

## Results

### Overview of Study cohorts

This study includes 3 large real-world patient clinical databases: (1) MarketScan Database which contains records from approximately 250 million patients from 2009–2022. (2) UMPH which includes approximately six million patients from 2006-2025. (3) OneFlorida+ Clinical Research Network which encompasses around 26 million patients from 72 hospitals across Florida, Alabama, Georgia, and Arkansas from 2013–2024. We employed 1:5 matching for PANC patients and controls, based on sex, age, and geographic location. As shown in Table 1, we identified 113,518, 8,257 and 10,500 PANC individuals in MartketScan, UMPH and OneFlorida+ respectively, among which the males account for 50.8%, 53.3% and 52.7%, similar among the datasets. The highest proportion of diagnoses is observed between ages 55 and 65, 65-70 and 65-75 (Fig. 3A, 3C and 3E). For MarketScan, most PANC patients exhibit trajectory lengths of 0–6 years with 50–400 accumulated diagnosis records (Fig. 3B), reflecting intensive healthcare utilization and multiple comorbidities in the years preceding diagnosis. Compared to the MarketScan database, PANC patients in UMPH generally have shorter trajectory lengths with most of them spanning 2 years or less, reflecting the high volume of referred high-risk patients in a research hospital (Fig. 3D). The OneFlorida+ cohort showed that PANC patients had trajectory lengths of ≤4 years (Fig. 3F) and accumulated a median of 96 diagnosis records (Table 1). In contrast, non-PANC patients generally have shorter trajectories in all three datasets, consistent with lower healthcare utilization.

**Fig 3.**
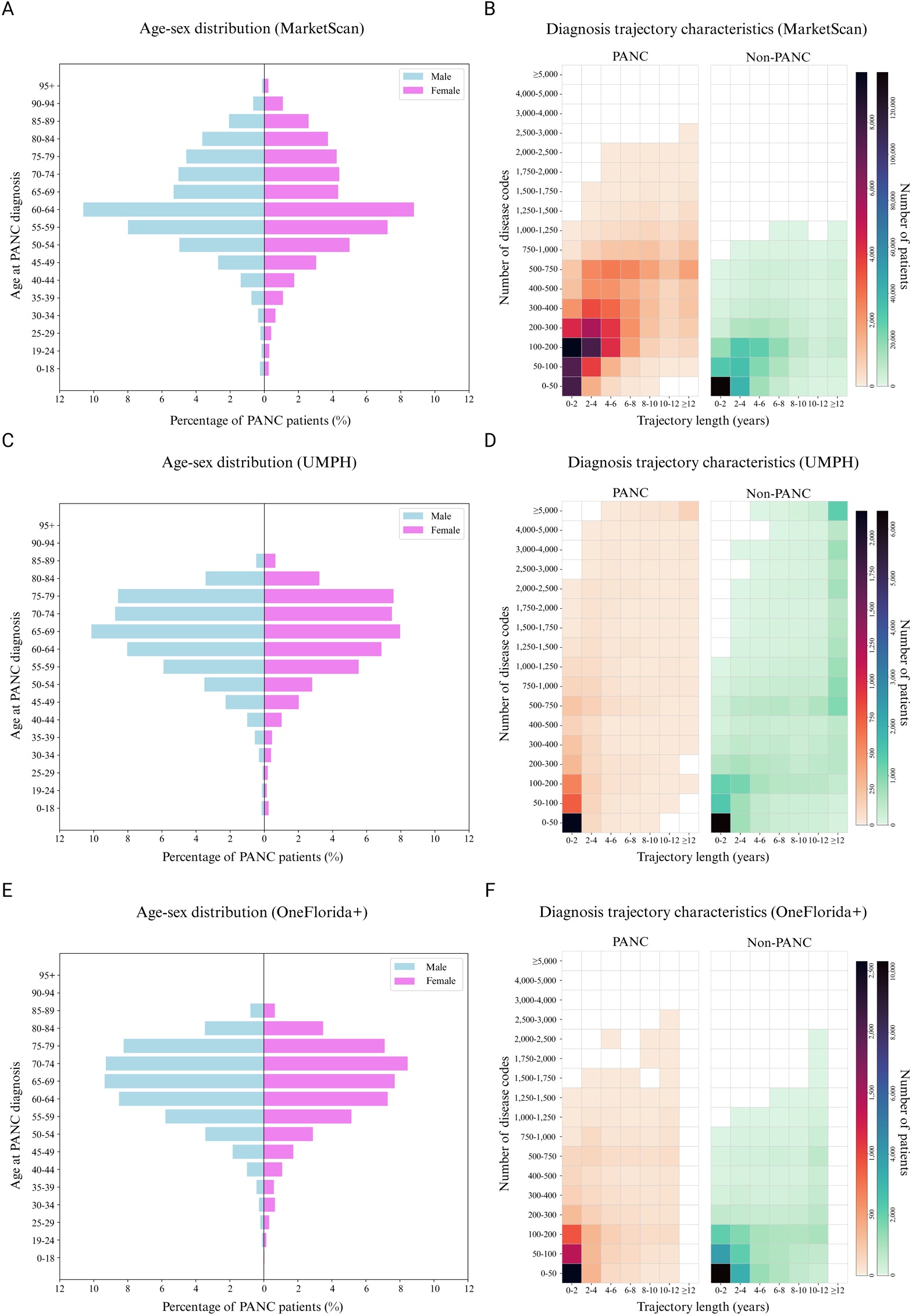
Characteristics of the MarketScan, UMPH and OneFlorida+ cohorts. Panels A,C, and E show the age-sex distribution of pancreatic cancer patients on MarketScan (Panel A), UMPH (Panel C) and OneFlorida+ (Panel E) cohorts, respectively. Panels B,D, and F show heatmaps of the joint distribution of diagnosis trajectory length and the number of distinct disease codes for PANC and non-PANC patients in the MarketScan (Panel B), UMPH (Panel D), and OneFlorida+ (Panel F) cohorts. Color intensity reflects the number of patients in each bin of age group, with darker shades indicating larger patient counts. UMPH: University of Michigan Precision Health; PANC: pancreatic cancer.

**Table 1:** Summary of patient statistics in the three datasets of this study.

| General dataset information | MarketScan | UMPH | OneFlorida+ |
| --- | --- | --- | --- |
| Dataset timeline | 2009-2022 | 2006-2025 | 2013-2024 |
| Total n patients | 680,821 | 48,318 | 54,853 |
| Male (%) | 346,084(50.8%) | 25,751(53.3%) | 27,596 (50.3%) |
| Female (%) | 334,737(49.2%) | 22,567(46.7%) | 27,257 (49.7%) |
| Median n disease codes per patient | 101 | 298 | 77 |
| Median length of trajectory in years | 3 | 5 | 2 |
| **Pancreatic cancer cohort information** |  |  |  |
| Total n patients | 113,518 | 8,257 | 10,500 |
| Male (%) | 57,713(50.8%) | 4,406(53.4%) | 5,537 (52.7%) |
| Female (%) | 55,805(49.2%) | 3,851(46.6%) | 4,963 (47.3%) |
| Median age at PC diagnosis | 62 | 67 | 67 |
| Median n disease codes per patient | 242 | 193 | 96 |
| Median length of trajectory in years | 3 | 1 | 1 |
| Smoker (%) | 24,965(22.0%) | 891(10.8%) | 4,319 (41.1%) |
| Alcohol user (%) | 5,520(4.9%) | 239(2.9%) | 693 (6.6%) |
| Illegal drug user (%) | 3,819(3.4%) | 328(4.0%) | 712 (6.8%) |

### Model Building on MarketScan Cohort

For each prediction interval setting, we obtained the model that achieved the minimum validation loss on the validation set, and then we evaluated its predictive ability on the MarketScan hold-out test set. As shown in Figs. 4A-B, the 6-month prediction window achieves the best performance, with an AUROC of 0.812 (95% CI, 0.807–0.818) and an AUPRG of 0.851 (95% CI, 0.843–0.860). The AUROC and AUPRG decrease as the prediction period gets longer, as expected. Still, the 60-month prediction window reaches the AUROC of 0.735 (95% CI, 0.724–0.745) and AUPRG of 0.629 (95% CI, 0.600–0.656). The shorter prediction window also has a larger training patient size, likely contributing to better model performance (Supplementary Table 6). To show where PANCDetect stands, we compared our model with CancerRiskNet^21^, with the same data preprocessing and the same training, validation, and testing cohorts. PANCDetect consistently outperforms CancerRiskNet across all prediction intervals (averaged AUC 0.633-0.765), particularly on long-term predictions over 36 and 60 months (Supplementary Fig. 2).

**Fig 4.**
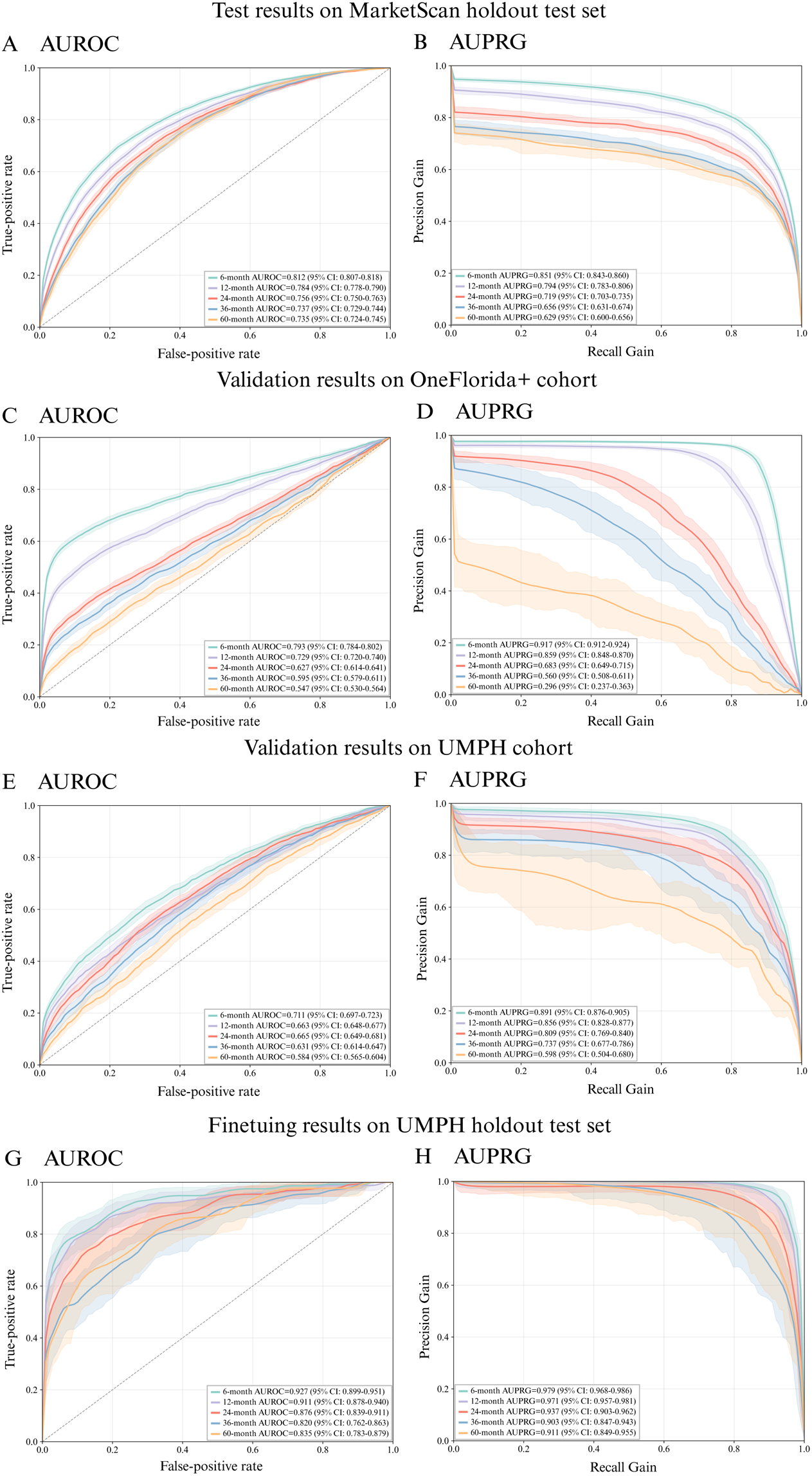
Model performance across MarketScan, UMPH, and OneFlorida+ cohorts. Panels A and B show results of the model trained on MarketScan training set tested on MarketScan hold-out test data; Panels C and D show OneFlorida+ external validation results using the model trained on MarketScan training data; Panels E and F show UMPH external validation results on the model trained on MarketScan training data; Panels G and H show fine-tuning results on UMPH hold-out data, after adding laboratory test data to the model built on MarketScan data. Panels A,C,E, and G: Results in AUROC; Panels B,D,F, and H: Results in AUPRG. Shaded areas represent 95% confidence intervals. AUROC: area under the receiver operating characteristic curve; AUPRG: area under the precision-recall-gain curve; UMPH: University of Michigan Precision Health; PANC: pancreatic cancer.

### Tests on the technical rigor of PANCDetect

We further evaluated several LLMs pretrained on biomedical corpora, such as BioGPT^17^, which is trained on PubMed abstracts, and GatorTron^28^, which is trained on large-scale de-identified clinical notes of University of Florida (UF) Health, PubMed articles, and Wikipedia. These domain-specific models were compared against the general-purpose embedding model OpenAI text-embedding-3-small, with predictive performance evaluated by AUROC and AUPRG on the test set. Experimental results show that the OpenAI text-embedding-3-small model achieves the performance that is comparable to, and in most cases better than, these specialized biomedical domain-specific models (Supplementary Table 7). This aligns with some of the existing literature showing that specialized models, whether trained from scratch or fine-tuned, often underperform compared to more general large-scale models across multiple tasks.^29,30^

We also performed another ablation study on multimodal fusion strategies by sequentially removing procedure and/or drug medication modalities (Supplementary Table 8). The model using diagnosis codes alone provided the baseline performance (AUROC of 0.779 and AUPRG of 0.810 for 6-month prediction). Adding procedures or drugs yielded consistent gains, with AUROC of 0.802 and AUPRG of 0.841, and AUROC of 0.801 and AUPRG of 0.828, for 6-month prediction, respectively. The best results were obtained when all three modalities were combined (AUROC of 0.812 and AUPRG of 0.851 for 6-month prediction). The results show that incorporating both procedure and drug information led to a significant improvement on predictive accuracy, revealing the complementary nature of these modalities.

### External Validation Results on UMPH and OneFlorida+ Cohorts

To further evaluate the generalizability of our models, we tested them on two additional cohorts from the OneFlorida+ research network and the UMPH (Fig. 4C-F). The model achieves comparable results on the OneFlorida+ cohort (Fig. 4C-D), particularly in short-term (6-12 months) prediction with AUROCs of 0.793 and 0.729, and AUPRGs of 0.917 and 0.859. The performance declines with longer exclusion intervals as expected, as fewer patients are remaining for training the model and the inherent difficulty for prediction over longer intervals. On the other hand, for the UMPH Cohort, the model shows decent but lower predictive AUROC scores, ranging from 0.711 at 6-month prediction to 0.584 at 60-month prediction (Fig. 4E). AUPRG, however, decreases as the exclusion interval lengthens, from 0.891 at 6-month prediction to 0.598 at 60 months (Fig. 4F).

### Fine-tuning results on the UMPH Cohort

The performance decline observed on the two EHR cohorts (particularly UMPH data), as compared to that of MarketScan hold-out data, is primarily attributable to the difference in EHR data structure as compared to the MarketScan claims-based data. This includes differences in coding systems, variable coverage, patterns of missingness,^31^ as well as the underlying patient populations, age distributions, and care-seeking behaviors, etc. Collectively, these factors contribute to input feature distribution shifts that limit the generalizability of the models trained on the MarketScan cohort originally. Therefore, it is necessary to fine-tune the model on the EHR cohort to improve adaptability and healthcare-specific predictive performance.

Thus, we fine-tuned the model previously trained from the MarketScan cohort, using the training subset of the UMPH hospital EHR data. We incorporated laboratory test results as an additional Lab modality. We kept the data processing pipeline and model architecture unchanged. As shown in Fig. 4G-H, the fine-tuning results on UM hold-out testing data drastically improved the model’s performance on UMPH test data. The AUROC reached 0.927 (95% CI, 0.899–0.951) and the AUPRG reached 0.979 (95% CI, 0.968–0.986) for the 6-month prediction window, whereas the 60-month prediction window achieved an AUROC of 0.835 (95% CI, 0.783–0.879) and an AUPRG of 0.911 (95% CI, 0.849–0.955).

### Top risk predictor for pancreatic cancer

Based on IG attribution analysis, we summarized the top ten risk factors identified across different prediction windows (Fig.5). In the MarketScan cohort, the model consistently highlighted type 2 diabetes mellitus and other diseases of pancreas (except acute pancreatitis) as the top 1 and top 2 most important risk factors across all prediction windows, respectively. Even though their absolute scores decrease with the increase of prediction interval, these abnormalities maintained high importance scores throughout, indicating that they are the most stable and significant signals preceding pancreatic cancer.^32–34^ Abdominal & pelvic pain and acute pancreatitis are additional top risk factors as well, though their importance decreases as the prediction interval increases. By 60-month prediction, abdominal & pelvic pain falls below the top 10 risk factors.

**Fig. 5.**
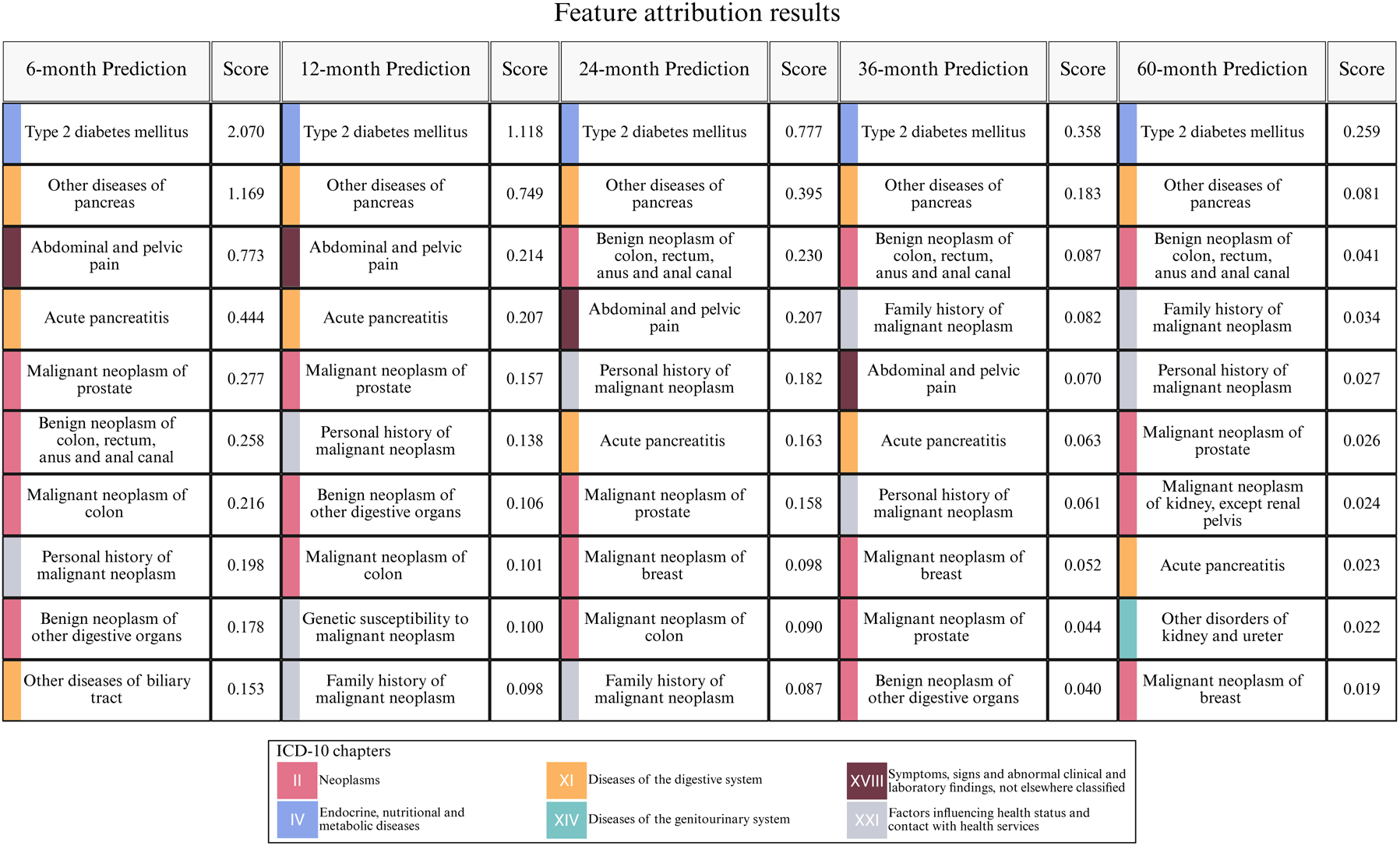
Attribution analysis of top EHR risk predictors for pancreatic cancer. Top 10 features from the model trained on MarketScan data. The top predictive clinical features across different prediction windows are ranked by the IG attribution scores.

In addition, personal or family history of malignant neoplasms repeatedly appear among the top 10 risk factors (Fig. 5), reflecting their importance in the preclinical phase of pancreatic cancer and potential comorbidity mechanism. In particular, neoplasm in different organs, such as colon and other GI tract organs, prostate and breast are also identified as important risk factors across different prediction windows, suggesting possible shared risk pathways or genetic susceptibility mechanisms with pancreatic cancer. Previous epidemiological studies have confirmed that these factors indeed elevate the risk of pancreatic cancer, thereby providing strong external evidence of our attribution result.^35–39^

## Discussion

This study leveraged the large-scale MarketScan database to develop a predictive model using multi-modal EHR information, including diagnoses, procedures and medications. Incorporating laboratory test data during fine-tuning on UM hospital validation cohort further improved model performance.

The strengths of this study lie in three areas. First, at the data level, the MarketScan database— covering a large and diverse population—provided a solid foundation for robust model development. We also conducted rigorous testing and validation on two regional hospital-based datasets, UMPH and OneFlorida+, to ensure the comprehensiveness of the study design and the reliability of the experimental results. Moreover, we show that inclusion of multimodal attributes in the EHR data can substantially improve the accuracy of cancer risk prediction through the ablation study. In particular, the inclusion of laboratory test results in hospital datasets significantly increased sensitivity and precision in the fine-tuned model. It is expected that with the inclusion of additional data modalities, such as unstructured clinical notes, the model performance will improve further. Second, at the methodological level, the proposed LLM-based embedding plus multimodal fusion framework also contributes to superior performance. This is demonstrated by better performance of PANCDetect in comparison to CancerRiskNet, using the same training, validation, and testing cohort. Interestingly, our embedding comparison revealed that representations derived from general LLMs outperformed those trained exclusively on biomedical text, underscoring the cross-domain transferability and robustness of general-purpose models. Third, at the study design level, the integration of IG-based interpretability revealed clinically meaningful drivers of predictions.

Nevertheless, several limitations remain. First, the generalizability of the model requires further improvement, as performance may decline when applied to external institutions or different populations. Second, discrepancies in coding systems across healthcare settings have not been resolved, posing challenges for real-world deployment and cross-system adaptation. Additionally, using ICD codes as diagnostic criteria is not perfectly accurate, as miscoding or incomplete documentation may introduce noise into the data. Thirdly, longer exclusion intervals tend to exclude a much larger number of patients and truncate the available trajectories, resulting in shorter and potentially less informative input sequences. This increases the difficulty of the prediction task over a long period of time. Federated learning frameworks could be utilized to facilitate more powerful long-term risk prediction, while preserving patient privacy, thereby improving generalizability^40^. Lastly, future efforts should focus on testing PANCDetect in prospective studies, before being deployed in real-world clinical environments for clinical decision support.

In conclusion, we present a multimodal disease risk prediction framework based on large-scale EHR data and applied it to pancreatic cancers, which demonstrated superior performance and interpretability compared with existing methods. Our model shows promise for potential applications in high-risk pancreatic cancer screening, early diagnosis, and personalized interventions. Ultimately, it may improve pancreatic cancer survival outcomes while reducing healthcare costs.

## Data Availability

This study analyzed de-identified EHR and claims data from three sources: the MarketScan Research databases (licensed from Merative), the University of Michigan Precision Health (UMPH), and the OneFlorida+ Clinical Research Network. Due to licensing restrictions, institutional regulations, and patient privacy considerations, these data are not publicly available. Access to MarketScan requires a data use agreement and paid license from Merative. Access to UMPH and OneFlorida+ data may be provided to qualified researchers upon reasonable request to the corresponding author and is subject to approval by the respective institutional review boards and data custodians.

## Acknowledgement

We would like to thank Dr. Anirban Maitra for his suggestion and encouragement to pursue this research, Patrick Brady and Institute for HealthCare Policy and Innovation (IHPI) of University of Michigan for helping us to access the MarketScan data. LXG would like to dedicate this work to her father Jianwen Xia, who passed away from pancreatic cancer on 09/28/2024.

## Author Contribution

LXG envisioned and supervised this project. ZJ, XG and ZW built and tested the model, conducted the analysis and wrote the manuscript. QY and RY conducted validation analysis using FloridaOne+ cohort. XY and XZ helped with analysis. XY helped to access UMPH data. All authors have read, revised and approved the final manuscript.

## Reference

1. Al-Shaheri, F. N. et al. Blood biomarkers for differential diagnosis and early detection of pancreatic cancer. Cancer Treat. Rev. 96, 102193 (2021).

2. Pereira, S. P. et al. Early detection of pancreatic cancer. Lancet Gastroenterol. Hepatol. 5, 698–710 (2020).

3. Huang, J. et al. Worldwide Burden of, Risk Factors for, and Trends in Pancreatic Cancer. Gastroenterology 160, 744–754 (2021).

4. Yang, J. et al. Early screening and diagnosis strategies of pancreatic cancer: a comprehensive review. Cancer Commun. 41, 1257–1274 (2021).

5. Søreide, K., Ismail, W., Roalsø, M., Ghotbi, J. & Zaharia, C. Early Diagnosis of Pancreatic Cancer: Clinical Premonitions, Timely Precursor Detection and Increased Curative-Intent Surgery. Cancer Control 30, 10732748231154711 (2023).

6. Elbanna, K. Y., Jang, H.-J. & Kim, T. K. Imaging diagnosis and staging of pancreatic ductal adenocarcinoma: a comprehensive review. Insights Imaging 11, 58 (2020).

7. Wittram, R., Kreis, L., König, H.-H. & Brettschneider, C. Economic evaluations of early detection strategies for pancreatic cancer: a systematic review. Eur. J. Health Econ. (2025).

8. Mishra, A. K., Chong, B., Arunachalam, S. P., Oberg, A. L. & Majumder, S. Machine Learning Models for Pancreatic Cancer Risk Prediction Using Electronic Health Record Data—A Systematic Review and Assessment. Am. J. Gastroenterol. 119, 1466–1482 (2024).

9. Guo, L. L. et al. A multi-center study on the adaptability of a shared foundation model for electronic health records. Npj Digit. Med. 7, 171 (2024).

10. Xian, S. et al. Transformer patient embedding using electronic health records enables patient stratification and progression analysis. Npj Digit. Med. 8, 521 (2025).

11. Zhang, S., Cornet, R. & Benis, N. Cross-Standard Health Data Harmonization using Semantics of Data Elements. Sci. Data 11, 1407 (2024).

12. Li, Y. et al. BEHRT: Transformer for Electronic Health Records. Sci. Rep. 10, 7155 (2020).

13. Li, Y. et al. Hi-BEHRT: Hierarchical Transformer-Based Model for Accurate Prediction of Clinical Events Using Multimodal Longitudinal Electronic Health Records. IEEE J. Biomed. Health Inform. 27, 1106–1117 (2023).

14. Rasmy, L., Xiang, Y., Xie, Z., Tao, C. & Zhi, D. Med-BERT: pretrained contextualized embeddings on large-scale structured electronic health records for disease prediction. Npj Digit. Med. 4, 86 (2021).

15. Su, X. et al. Multimodal Medical Code Tokenizer. Preprint at 10.48550/ARXIV.2502.04397 (2025).

16. Lee, S. A., Wu, A. & Chiang, J. N. Clinical ModernBERT: An efficient and long context encoder for biomedical text. Preprint at 10.48550/ARXIV.2504.03964 (2025).

17. Luo, R. et al. BioGPT: Generative Pre-trained Transformer for Biomedical Text Generation and Mining. Brief. Bioinform. 23, bbac409 (2022).

18. Jiang, L. Y. et al. Health system-scale language models are all-purpose prediction engines. Nature 619, 357–362 (2023).

19. Jia, K. et al. A pancreatic cancer risk prediction model (Prism) developed and validated on large-scale US clinical data. eBioMedicine 98, 104888 (2023).

20. Appelbaum, L. et al. Development and validation of a pancreatic cancer risk model for the general population using electronic health records: An observational study. Eur. J. Cancer 143, 19–30 (2021).

21. Placido, D. et al. A deep learning algorithm to predict risk of pancreatic cancer from disease trajectories. Nat. Med. 29, 1113–1122 (2023).

22. Park, J. et al. Enhancing EHR-based pancreatic cancer prediction with LLM-derived embeddings. Npj Digit. Med. 8, 465 (2025).

23. Butler, A. M., Nickel, K. B., Overman, R. A. & Brookhart, M. A. IBM MarketScan Research Databases. in Databases for Pharmacoepidemiological Research (eds Sturkenboom, M. & Schink, T.) 243–251 (Springer International Publishing, Cham, 2021). doi:10.1007/978-3-030-51455-6_20.

24. Zawistowski, M. et al. The Michigan Genomics Initiative: A biobank linking genotypes and electronic clinical records in Michigan Medicine patients. Cell Genomics 3, 100257 (2023).

25. Shenkman, E. et al. OneFlorida Clinical Research Consortium: Linking a Clinical and Translational Science Institute With a Community-Based Distributive Medical Education Model. Acad. Med. 93, 451–455 (2018).

26. Flach, P. & Kull, M. Precision-Recall-Gain Curves: PR Analysis Done Right. In Proc. 28th Conference on Neural Information Processing Systems (NeurIPS) (2015).

27. Sundararajan, M., Taly, A. & Yan, Q. Axiomatic Attribution for Deep Networks. In Proc. 34th International Conference on Machine Learning (ICML) (2017).

28. Yang, X. et al. A Large Clinical Language Model to Unlock Patient Information from Unstructured Electronic Health Records. Npj Digit. Med. 5, 194 (2022).

29. Excoffier, J.-B. et al. Generalist embedding models are better at short-context clinical semantic search than specialized embedding models. Preprint at https://doi.org/10.48550/ARXIV.2401.01943 (2024).

30. Hegselmann, S. et al. Large Language Models are Powerful Electronic Health Record Encoders. Preprint at 10.48550/ARXIV.2502.17403 (2025).

31. Laws, M. B., Michaud, J., Shield, R., McQuade, W. & Wilson, I. B. Comparison of Electronic Health Record–Based and Claims-Based Diabetes Care Quality Measures: Causes of Discrepancies. Health Serv. Res. 53, 2988–3006 (2018).

32. Cho, J., Scragg, R. & Petrov, M. S. Postpancreatitis Diabetes Confers Higher Risk for Pancreatic Cancer Than Type 2 Diabetes: Results From a Nationwide Cancer Registry. Diabetes Care 43, 2106–2112 (2020).

33. Huxley, R., Ansary-Moghaddam, A., Berrington de González, A., Barzi, F. & Woodward, M. Type-II diabetes and pancreatic cancer: a meta-analysis of 36 studies. Br. J. Cancer 92, 2076–2083 (2005).

34. Lai, S.-W. & Liao, K.-F. Pancreatitis, Type 2 Diabetes Mellitus, and Pancreatic Cancer (Diabetes Metab J 2025;49:252-63). Diabetes Metab. J. 49, 518–519 (2025).

35. Price, S. J., Gibson, N., Hamilton, W. T., King, A. & Shephard, E. A. Intra-abdominal cancer risk with abdominal pain: a prospective cohort primary care study. Br. J. Gen. Pract. J. R. Coll. Gen. Pract. 72, e361–e368 (2022).

36. Matsubayashi, H. et al. Surveillance of Individuals with a Family History of Pancreatic Cancer and Inherited Cancer Syndromes: A Strategy for Detecting Early Pancreatic Cancers. Diagn. Basel Switz. 9, 169 (2019).

37. Seppälä, T. T., Burkhart, R. A. & Katona, B. W. Hereditary colorectal, gastric, and pancreatic cancer: comprehensive review. BJS Open 7, zrad023 (2023).

38. Mocci, E. et al. Risk of pancreatic cancer in breast cancer families from the breast cancer family registry. Cancer Epidemiol. Biomark. Prev. Publ. Am. Assoc. Cancer Res. Cosponsored Am. Soc. Prev. Oncol. 22, 803–811 (2013).

39. Jacobs, E. J. et al. Family history of cancer and risk of pancreatic cancer: a pooled analysis from the Pancreatic Cancer Cohort Consortium (PanScan). Int. J. Cancer 127, 1421–1428 (2010).

40. Sadilek, A. et al. Privacy-first health research with federated learning. Npj Digit. Med. 4, 132 (2021).

