## Supplementary Appendix for "PANCDetect: Early Detection of Pancreatic Cancer from Multimodal EHR data with LLM Embeddings"

**Integrated gradient (IG)[1] implementation details:**

The IG along the $i^{th}$ dimension is defined as the following:

$$IG_i(x) := (x_i - x_i') \times \int_0^1 \frac{\partial F(x' + \alpha(x - x'))}{\partial x_i} d\alpha$$

where $x$ is the actual input, $x'$ is the baseline (often chosen as zeros or some "neutral" input), $F$ is the prediction function (e.g., the model's risk score for pancreatic cancer), $\alpha$ is an interpolation parameter that smoothly moves from baseline ($\alpha$=0) to actual input ($\alpha$=1).

For implementation, the integral can't be computed analytically for deep models, we approximate the integration via the summation with steps m=30 as the following:

$$IG_i^{approx}(x) := (x_i - x_i') \times \sum_{k=1}^{m} \frac{\partial F(x' + \frac{k}{m}(x - x'))}{\partial x_i} \times \frac{1}{m}$$

So practically, IG is "average gradient × input difference."

**Supplementary Fig. 1**: Age and sex distributions of PANC patients (left) and matched non-PANC controls (right) in the MarketScan cohort. A 1:5 case–control matching strategy based on sex, age, and geographic location ensured consistent demographic ratios between cases and controls. We kept the same matching strategy for UMPH and OneFlorida+ cohorts too.

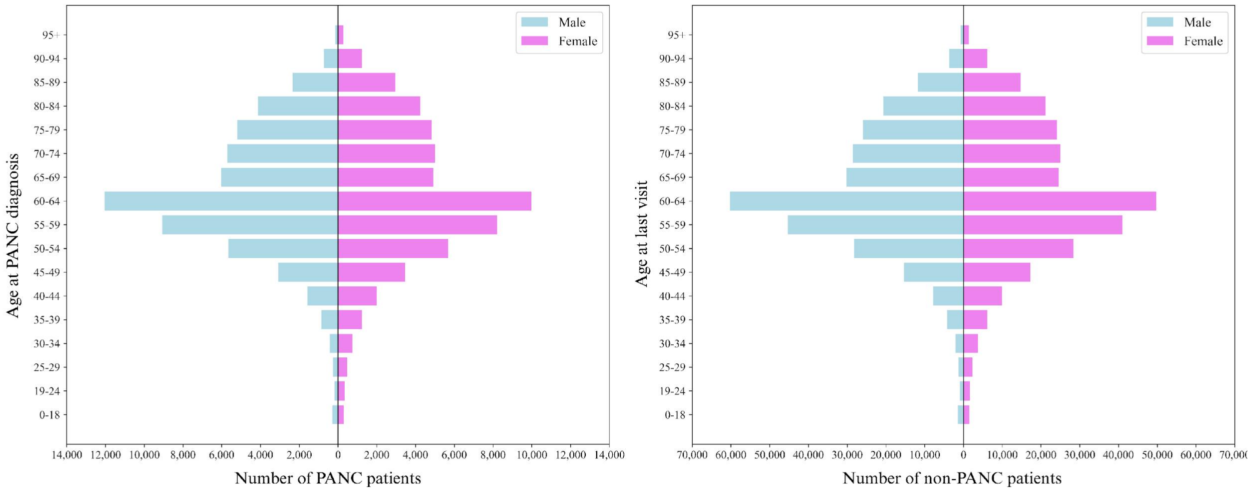

**Supplementary Fig. 2**: Comparative performance of PANCDetect and CancerRiskNet under the same prediction interval setting. PANCDetect achieved higher AUROC (A) and AUPRG (B) scores across all intervals, with particularly strong gains in long-term predictions (36- and 60-month). CancerRiskNet showed lower discrimination and precision–recall gain, especially at extended horizons. Shaded areas represent 95% confidence intervals.

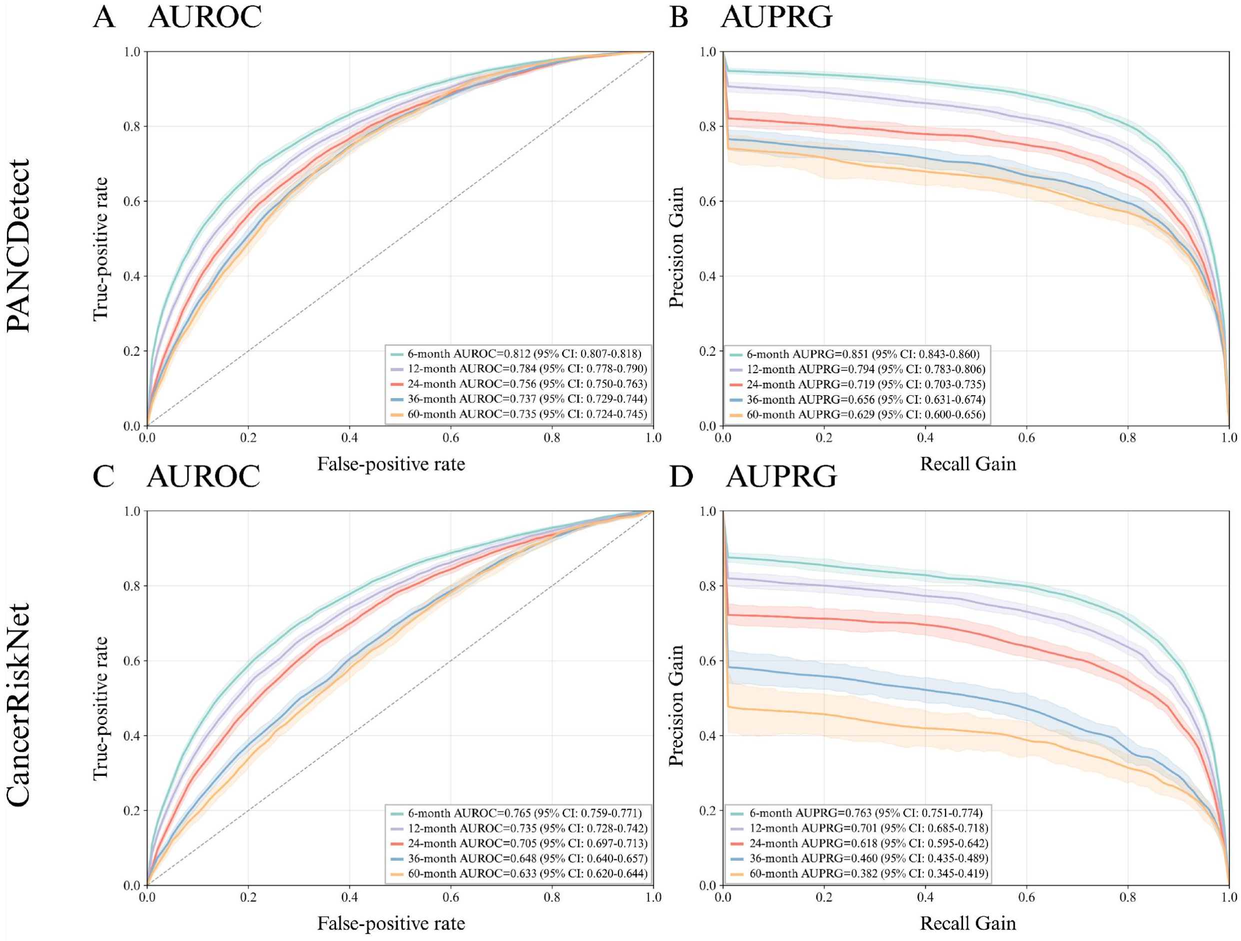

**Supplementary Table 1**: ICD 9/10 Code for pancreatic cancer

| ICD 9 Code | ICD 10 Code |
|---|---|
| 157: Malignant neoplasm of pancreas | C25: Malignant neoplasm of pancreas |
| 157.0: Malignant neoplasm of head of pancreas | C25.0: Malignant neoplasm of head of pancreas |
| 157.1: Malignant neoplasm of body of pancreas | C25.1: Malignant neoplasm of body of pancreas |
| 157.2: Malignant neoplasm of tail of pancreas | C25.2: Malignant neoplasm of tail of pancreas |
| 157.3: Malignant neoplasm of pancreatic duct | C25.3: Malignant neoplasm of pancreatic duct |
| 157.4: Malignant neoplasm of islets of langerhans | C25.4: Malignant neoplasm of endocrine pancreas |
| 157.8: Malignant neoplasm of other specified sites of pancreas | C25.7: Malignant neoplasm of other parts of pancreas |
| 157.9: Malignant neoplasm of pancreas, part unspecified | C25.8: Malignant neoplasm of overlapping sites of pancreas |
| | C25.9: Malignant neoplasm of pancreas, unspecified |

**Supplementary Table 2**: Other evaluation metrics performance on MarketScan test set.

| | **6-month** | **12-month** | **24-month** | **36-month** | **60-month** |
|---|---|---|---|---|---|
| **Precision** | 0.626 (95% CI: 0.612-0.638) | 0.634 (95% CI: 0.616-0.653) | 0.502 (95% CI: 0.484-0.525) | 0.575 (95% CI: 0.537-0.607) | 0.433 (95% CI: 0.405-0.455) |
| **Recall** | 0.360 (95% CI: 0.350-0.369) | 0.235 (95% CI: 0.225-0.246) | 0.258 (95% CI: 0.246-0.270) | 0.106 (95% CI: 0.097-0.116) | 0.306 (95% CI: 0.290-0.324) |
| **F1 Score** | 0.458 (95% CI: 0.445-0.467) | 0.343 (95% CI: 0.332-0.357) | 0.341 (95% CI: 0.327-0.355) | 0.180 (95% CI: 0.165-0.194) | 0.359 (95% CI: 0.340-0.377) |
| **Specificity** | 0.955 (95% CI: 0.953-0.957) | 0.970 (95% CI: 0.969-0.972) | 0.944 (95% CI: 0.941-0.947) | 0.982 (95% CI: 0.980-0.984) | 0.901 (95% CI: 0.895-0.907) |

**Supplementary Table 3**: Other evaluation metrics performance on UMPH cohort (external validation).

| | **6-month** | **12-month** | **24-month** | **36-month** | **60-month** |
|---|---|---|---|---|---|
| **Precision** | 0.200 (95% CI: 0.189-0.214) | 0.212 (95% CI: 0.196-0.233) | 0.154 (95% CI: 0.137-0.169) | 0.132 (95% CI: 0.116-0.152) | 0.063 (95% CI: 0.055-0.071) |
| **Recall** | 0.373 (95% CI: 0.350-0.391) | 0.244 (95% CI: 0.224-0.262) | 0.225 (95% CI: 0.201-0.245) | 0.130 (95% CI: 0.110-0.149) | 0.223 (95% CI: 0.197-0.254) |
| **F1 Score** | 0.260 (95% CI: 0.244-0.275) | 0.227 (95% CI: 0.211-0.245) | 0.183 (95% CI: 0.166-0.199) | 0.131 (95% CI: 0.113-0.152) | 0.098 (95% CI: 0.086-0.111) |
| **Specificity** | 0.900 (95% CI: 0.897-0.904) | 0.948 (95% CI: 0.945-0.950) | 0.938 (95% CI: 0.935-0.941) | 0.961 (95% CI: 0.959-0.964) | 0.875 (95% CI: 0.871-0.879) |

**Supplementary Table 4**: Other evaluation metrics performance on OneFlorida+ cohort (external validation).

| | **6-month** | **12-month** | **24-month** | **36-month** | **60-month** |
|---|---|---|---|---|---|
| **Precision** | 0.482 (95% CI: 0.467-0.493) | 0.502 (95% CI: 0.484-0.521) | 0.351 (95% CI: 0.331-0.371) | 0.447 (95% CI: 0.409-0.480) | 0.272 (95% CI: 0.239-0.303) |
| **Recall** | 0.618 (95% CI: 0.600-0.634) | 0.448 (95% CI: 0.429-0.464) | 0.319 (95% CI: 0.299-0.335) | 0.203 (95% CI: 0.185-0.224) | 0.170 (95% CI: 0.147-0.196) |
| **F1 Score** | 0.541 (95% CI: 0.527-0.553) | 0.473 (95% CI: 0.457-0.488) | 0.334 (95% CI: 0.317-0.351) | 0.279 (95% CI: 0.253-0.302) | 0.209 (95% CI: 0.182-0.234) |
| **Specificity** | 0.891 (95% CI: 0.887-0.896) | 0.927 (95% CI: 0.924-0.931) | 0.898 (95% CI: 0.894-0.903) | 0.955 (95% CI: 0.950-0.958) | 0.915 (95% CI: 0.908-0.922) |

**Supplementary Table 5**: Other evaluation metrics performance on UMPH hold-out test cohort (finetuning).

| | **6-month** | **12-month** | **24-month** | **36-month** | **60-month** |
|---|---|---|---|---|---|
| **Precision** | 0.799 (95% CI: 0.736-0.862) | 0.809 (95% CI: 0.750-0.872) | 0.736 (95% CI: 0.649-0.819) | 0.807 (95% CI: 0.695-0.906) | 0.673 (95% CI: 0.541-0.780) |
| **Recall** | 0.686 (95% CI: 0.610-0.756) | 0.654 (95% CI: 0.583-0.767) | 0.565 (95% CI: 0.496-0.641) | 0.400 (95% CI: 0.333-0.481) | 0.438 (95% CI: 0.337-0.547) |
| **F1 Score** | 0.738 (95% CI: 0.679-0.793) | 0.724 (95% CI: 0.664-0.775) | 0.639 (95% CI: 0.575-0.700) | 0.535 (95% CI: 0.457-0.617) | 0.530 (95% CI: 0.417-0.632) |
| **Specificity** | 0.969 (95% CI: 0.958-0.979) | 0.967 (95% CI: 0.956-0.980) | 0.957 (95% CI: 0.941-0.973) | 0.980 (95% CI: 0.968-0.991) | 0.957 (95% CI: 0.939-0.973) |

**Supplementary Table 6:** Cohort size settings for model training, validation, and finetuning.

| | **Case/Control** | | | | |
|---|---|---|---|---|---|
| **Cohorts** | **6-month** | **12-month** | **24-month** | **36-month** | **60-month** |
| **MarketScan (Model building)** | | | | | |
| Training | 59,106/283,37 | 50,499/238,61 | 38,387/175,82 | 29,022/129,31 | 16,557/70,00 |
| Validation | 7,485/35,324 | 6,393/29,747 | 4,796/21,980 | 3,652/16,140 | 2,111/8,709 |
| Hold-out testing | 7,429/35,381 | 6,401/29,739 | 4,832/21,945 | 3,720/16,073 | 2144/8,676 |
| **OneFlorida+ (External validation)** | | | | | |
| Testing | 3,878/23,743 | 3,074/18,784 | 2,297/13,307 | 1,815/10,051 | 1,187/6,309 |
| **UMPH (External validation)** | | | | | |
| Testing | 1,758/26,326 | 1,585/27,416 | 1,334/26,728 | 1,129/24,829 | 804/21,252 |
| **UMPH (Finetuning)** | | | | | |
| Training | 1,422/6,822 | 1,262/6,166 | 1,072/5,192 | 902/4,416 | 633/3,146 |
| Validation | 180/851 | 161/768 | 124/659 | 112/553 | 91/381 |
| Testing | 156/875 | 162/767 | 138/646 | 115/550 | 80/393 |

**Supplementary Table 7**: Ablation study of different embedding generation methods.

| **Model name & embedding dimension** | **6-month prediction** | | **60-month prediction** | |
|---|---|---|---|---|
| | **AUROC** | **AUPRG** | **AUROC** | **AUPRG** |
| **BioGPT (1024)** | 0.801 (95% CI: 0.793-0.805) | 0.837 (95% CI: 0.829-0.846) | 0.710 (95% CI: 0.698-0.721) | 0.578 (95% CI: 0.547-0.607) |
| **GatorTron (1024)** | 0.812 (95% CI: 0.806-0.817) | **0.853** (95% CI: 0.836-0.861) | 0.729 (95% CI: 0.718-0.739) | 0.623 (95% CI: 0.595-0.651) |
| **Trained from scratch (128)** | 0.772 (95% CI: 0.764-0.777) | 0.802 (95% CI: 0.791-0.812) | 0.682 (95% CI: 0.670-0.694) | 0.520 (95% CI: 0.484-0.553) |
| **OpenAI text-embedding-3-small (1536)** | **0.812** (95% CI: 0.807-0.818) | 0.851 (95% CI: 0.843-0.860) | **0.735** (95% CI: 0.724-0.745) | **0.629** (95% CI: 0.600-0.656) |

**Supplementary Table 8**: Ablation study of different modalities used for training.

| **Modalities** | **6-month prediction** | | **60-month prediction** | |
|---|---|---|---|---|
| | **AUROC** | **AUPRG** | **AUROC** | **AUPRG** |
| **Diagnosis** | 0.779 (95% CI: 0.773-0.784) | 0.810 (95% CI: 0.799-0.819) | 0.708 (95% CI: 0.698-0.719) | 0.558 (95% CI: 0.527-0.586) |
| **Diagnosis + Procedure** | 0.802 (95% CI: 0.796-0.807) | 0.841 (95% CI: 0.832-0.850) | 0.721 (95% CI: 0.711-0.732) | 0.603 (95% CI: 0.575-0.627) |
| **Diagnosis + Drug** | 0.801 (95% CI: 0.794-0.806) | 0.828 (95% CI: 0.818-0.838) | 0.729 (95% CI: 0.718-0.739) | 0.618 (95% CI: 0.590-0.643) |
| **Diagnosis + Procedure + Drug** | **0.812** (95% CI: 0.807-0.818) | **0.851** (95% CI: 0.843-0.860) | **0.735** (95% CI: 0.724-0.745) | **0.629** (95% CI: 0.600-0.656) |

### Reference


1. Sundararajan, M., Taly, A. & Yan, Q. Axiomatic Attribution for Deep Networks. *In Proc. 34th International Conference on Machine Learning (ICML)* (2017).
